# Epigenome-wide contributions to individual differences in childhood phenotypes: A GREML approach

**DOI:** 10.1101/2021.06.24.21259449

**Authors:** Alexander Neumann, Jean-Baptiste Pingault, Janine F. Felix, Vincent W. V. Jaddoe, Henning Tiemeier, Charlotte Cecil, Esther Walton

## Abstract

**Background:** DNA methylation is an epigenetic mechanism involved in human development. Numerous epigenome-wide association studies (EWAS) have investigated the associations of DNA methylation at single CpG sites with childhood outcomes. However, the overall contribution of DNA methylation across the genome (R^2^_Methylation_) towards childhood phenotypes is unknown. An estimate of R^2^_Methylation_ would provide context regarding the importance of DNA methylation explaining variance in health outcomes.

**Methods:** We estimated the variance explained by epigenome-wide cord blood methylation (R^2^_Methylation_) for five childhood phenotypes: gestational age, birth weight, and body mass index (BMI), IQ and ADHD symptoms at school age. We adapted a genome-based restricted maximum likelihood (GREML) approach with cross-validation (CV) to DNA methylation data and applied it in two population-based birth cohorts: ALSPAC (n=775) and Generation R (n=1382).

**Results:** Using information from >470,000 autosomal probes we estimated that DNA methylation at birth explains 45% (SD_CV_ = 0.07) of gestational age variance and 16% (SD_CV_ = 0.05) of birth weight variance. The R^2^_Methylation_ estimates for BMI, IQ and ADHD symptoms at school age estimates were near 0% across almost all cross-validation iterations.

**Conclusions:** The results suggest that cord blood methylation explains a moderate to large degree of variance in gestational age and birth weight, in line with the success of previous EWAS in identifying numerous CpG sites associated with these phenotypes. In contrast, we could not obtain a reliable estimate for school-age BMI, IQ and ADHD symptoms. This may reflect a null bias due to insufficient sample size to detect variance explained in more weakly associated phenotypes, although the true R^2^_Methylation_ for these phenotypes is likely below that of gestational age and birth weight when using DNA methylation at birth.

## Background

DNA methylation (DNAm) is an epigenetic process, which involves the attachment of a methyl group to cytosine bases, typically in the context of a cytosine-phosphate-guanine dinucleotide (CpG) site. The methylation status of a CpG site can have an impact on gene expression and downstream phenotypes (1). In turn, methylation levels are determined by genetics, environment and stochastic processes (2,3). DNAm could therefore function as mediator of many genetic and environmental determinants of human development, functioning and pathology. A common research design to query the role of DNAm in these processes is an epigenome-wide association study (EWAS). As a large number of CpG sites are tested, to reliably identify relevant CpG sites, either large samples or big effect sizes are required, which for most traits or CpG sites are not available or unlikely (4).

However, analogous to lessons learned from genome-wide association studies, no matter the number of genome-wide significant CpGs identified in an EWAS, whether it be 0 or thousands, there is always a possibility that more CpGs are associated with a predictor or outcome, but did not reach significance due to lack of power. Since an EWAS estimates the associations of single CpG probes, no conclusions can be drawn about the overall contribution of genome-wide DNAm towards a phenotype. Such an overall estimate of variance explained by genome-wide DNAm (R^2^_Methylation_) would be highly informative for several reasons: 1. R^2^_Methylation_ would provide a picture of how relevant DNAm levels are to an outcome, either as causal determinant or predictor. 2. R^2^_Methylation_ would provide an upper limit of how much the combined effects of CpG sites identified by an EWAS (e.g. poly-epigenetic score) can explain. While estimates of R^2^_Methylation_ would be clearly useful, the best approach to derive them is less clear. One option is to adapt the genomic restricted maximum likelihood (GREML) (5) approach used in genetics.

In genetics, the analogous measure of R^2^_Methylation_ is the single nucleotide polymorphism heritability (SNP h^2^), i.e. the variance explained by all measured SNPs. A popular method to estimate SNP h^2^ is through a GREML analysis which consists of two steps: 1. The estimation of genetic relatedness values between participant pairs inferred from their similarity in measured SNP genotypes. 2. Estimating how well genetic relatedness predicts phenotypic similarity between participant pairs. While the GREML approach has been developed for genetic data, the analysis can be applied to any high dimensional data, such as genome-wide methylation data. First papers are now being published using GREML and alternative methods to estimate the variance explained by genome-wide DNAm. An early example is a study by Vazquez et al. (6), who used a Bayesian variant of a GREML model to predict breast cancer survival. The authors found that genome-wide DNAm is more predictive than the structural genome or traditional covariates alone, explaining 16.2% of variance. More recently, Zhang et al. (7) tested the validity of the GREML approach in methylation data using simulations and real data in a sample of adults. The authors estimated that concurrent blood DNAm levels explained 6.5% of the variance in BMI but were not associated with height, when controlling for genetic effects. In contrast, using a Bayesian approach not relying on similarity matrices, Banos et al. (8) estimated the proportion of BMI variance explained by concurrent DNAm to be 75.7% in adulthood. The CpG-level effects estimated by this model explained up to 30.8% in adult replications cohorts, but only 3.3%, 2.05% and 9.65% at birth, age 7 and age 15 respectively, with BMI and DNAm measured at the same time-points. The results suggest highly age specific effects depending on when both BMI and DNAm were measured.

As previous studies focused on DNAm and outcomes in adults, the variance of childhood outcomes explained by cord blood DNAm is unknown. In this study we aimed to use cord blood DNAm to estimate the R^2^_Methylation_ of five child outcomes, previously addressed in EWAS studies: gestational age and birth weight, as well as BMI, IQ and ADHD symptoms at school age. These outcomes were chosen because they represent childhood outcomes in different areas (general health, cognition and psychopathology). In addition, all of these have been studied in multi-center population-based EWAS before, allowing for a comparison between R^2^_Methylation_ measures and EWAS findings. Two of the phenotypes most robustly associated with DNAm in EWAS studies are gestational age and birth weight. For gestational age, 8899 CpGs have been found to be significantly associated in a previous EWAS at genome-wide significance (9). Prediction models based on these CpGs were able to explain 50-80% of the gestational age variance in an independent sample (10,11). In the case of birth weight, 914 sites were associated based on an EWAS meta-analysis in 8,825 children (12). Cord blood has also the potential to predict later development, e.g. nine CpG sites were associated with ADHD symptoms in school-age according to a recent EWAS in 2,477 children (13) and one CpG site predicted BMI in late childhood (n=4133) (14). In contrast, no genome-wide significant sites in cord blood were identified for BMI in early childhood (14) nor IQ in school-age (n=3798) (15). While the variance explained by specific sets of CpGs is known for some childhood outcomes, the genome-wide contribution has not been studied before. The aim of this study is to estimate the genome-wide contribution of cord blood DNA to various childhood outcomes.

## Methods

### Participants

Participants for this study were drawn from two European population-based birth cohorts: The ALSPAC Study and the Generation R study. ALSPAC had recruited 15,454 women with an expected delivery date between April 1991 and December 1992, who were living in the former English county Avon, resulting in 15,589 foetuses. Of these 14,901 were alive at 1 year of age. The development of their children was subsequently studied at multiple assessment waves. Cord blood DNAm was assessed for 1,018 children. To avoid potential biases arising from shared family environment or population stratification, only one sibling per family was included in the analyses sample, as well as only children whose parents reported white ethnicity (analysis n=775). Full cohort descriptions have been published previously (16,17). Please note that the study website contains details of all the data that is available through a fully searchable data dictionary and variable search tool (http://www.bristol.ac.uk/alspac/researchers/our-data/). Ethical approval for the study was obtained from the ALSPAC Ethics and Law Committee and the Local Research Ethics Committees. Consent for biological samples has been collected in accordance with the Human Tissue Act (2004). Informed consent for the use of data collected via questionnaires and clinics was obtained from participants following the recommendations of the ALSPAC Ethics and Law Committee at the time.

Generation R invited all pregnant women living in the city of Rotterdam, the Netherlands, with an expected delivery date between April 2002 and January 2006 to participate in the study, of which 9,778 were enrolled. Cord blood DNA methylation was assessed in a subgroup of 1396 children with parents of reported European national origin. After exclusion of siblings (one of each pair excluded), 1382 participants remained in the analysis. Full study descriptions have been published previously (18), see also https://generationr.nl/researchers/ for more information. All parents gave informed consent for their children’s participation. The Generation R Study is conducted in accordance with the Declaration of Helsinki. Study protocols were approved by the Ethics Committee of Erasmus MC.

### Measures

#### DNA Methylation

DNAm was measured in cord blood at birth. Bisulfite conversion was performed with the EZ-96 DNAm kit (shallow) (Zymo Research Corporation, Irvine, USA). DNAm levels were then measured with the Illumina Infinium HumanMethylation450 BeadChip array (Illumina Inc., San Diego, USA). Preprocessing in ALSPAC was performed with the meffil package (19). Quality control check included mismatched genotypes, mismatched sex, incorrect relatedness, low concordance with other time points, extreme dye bias, and poor probe detection. In Generation R, pre-processing was performed with the CPACOR workflow (20). Quality control exclusion criteria included failed bisulfite conversion, hybridization or extension, sex mismatches and call rate <= 95%. Both cohorts were normalized using a combined dataset, using meffil functional normalization with ten control probe principal components and slide included as a random effect, see Mulder et al. (21) for further details. To lessen the influence of methylation outliers while retaining a consistent sample size, extreme values were winsorized. Per CpG site, DNAm levels exceeding three times the interquartile range above the third or below the first quartile (3*IQR criterion) were replaced by the maximum or minimum value, respectively, of the sample below the exclusion criterion. Only autosomal probes were considered in this study for consistent interpretation of effects between sexes. This resulted in 470,870 and 473,864 CpG probes in ALSPAC and Generation R, respectively, which were used for the computation of the methylation similarity matrix.

#### Outcomes and covariates

##### Birth outcomes

In ALSPAC, birthweight was recorded by healthcare professionals at the time of birth and extracted from birth records (12). Gestational age at delivery was also extracted from birth records. Obstetric practice and antenatal care at the time means that for most participants gestational age will have been estimated based on the last menstrual period, supplemented by ultrasound scans and paediatric/obstetric assessment of the newborn at birth.

In GenR, midwife and hospital registries were used to obtain information on birth weight. Gestational age was based on ultrasound examinations for mothers who enrolled in early or mid pregnancy, but based on last menstrual period for late pregnancy (22).

##### Childhood outcomes

In ALSPAC, measurements of height and weight, with the participant in light clothing and without shoes, were obtained at clinic visits when the children were seven years of age to calculate BMI. Non-verbal IQ at age 8 years was measured by the Wechsler Intelligence Scale for Children WISC-III UK (23). ADHD symptomatology was assessed via maternal ratings at age 7, with the Development and Well-Being Assessment interview (DAWBA) (24).

In Generation R, when children were 6.0 (SD=0.15) years old, children’s height and weight were measured at the research center without shoes or heavy clothing and used for the calculation of BMI (kg/m^2^). Non-verbal IQ was assessed at the same age using the Snijder-Oomen nonverbal intelligence test (25). ADHD symptoms were rated by a primary caregiver (90% mothers) using the Conners’ Parent Rating Scale-Revised (CPRS-R) questionnaire at age 8.1 (SD=0.15) (26).

##### Covariates

In ALSPAC, mothers were asked about their smoking during pregnancy, and these data were used to generate a binary variable of any smoking during pregnancy. Maternal education was collapsed into whether they had achieved a university degree or not. Across cohorts, white cell proportions were estimated with the Houseman method using the cord blood specific Bakulski reference (27). In Generation R, maternal age was obtained at enrollment. Maternal smoking was defined as either “Never smoked”, “Quit smoking in early pregnancy”, “Continued smoking during pregnancy”. Maternal education during pregnancy was categorized as “no education”, “primary education”, “secondary education first phase”, “secondary education second phase”, higher education first phase”, higher education second phase”. See Table 1 for descriptive statistics of all variables.

**Table 1:**
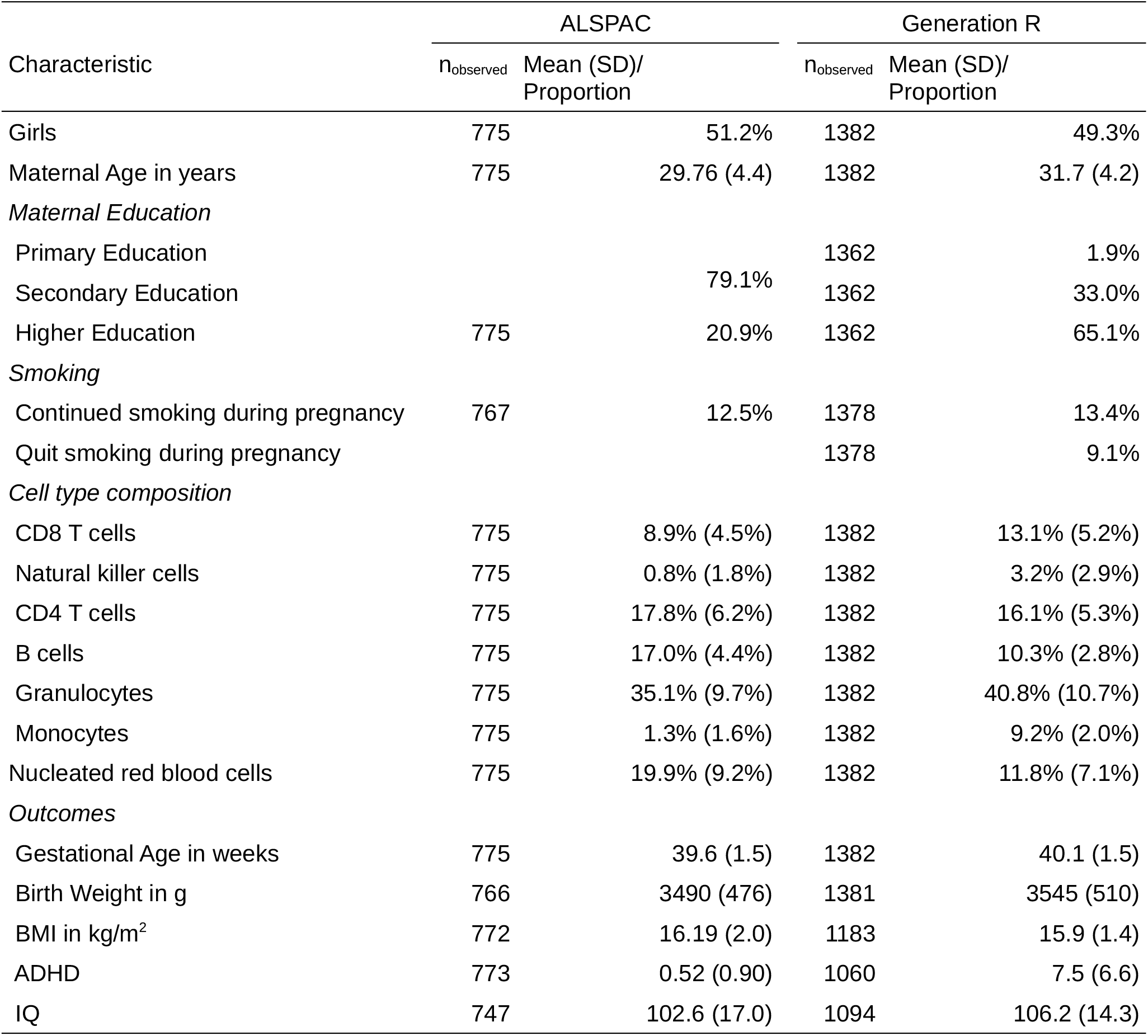
Participant Characteristics

### Statistical Analysis

We adapted the GREML approach to estimate R^2^_Methylation_. The GREML procedure consists of two steps: 1. Compute a genetic relatedness matrix (i.e. how genetically similar two individuals are based on SNP data), 2. Regress the outcome similarity between participants on the genetic relatedness (i.e. to establish whether greater genetic similarity between individuals relates to greater phenotypic similarity).

We refer to a methylation similarity matrix (M) as opposed to a genetic relatedness matrix (G). However, both M and G can be calculated with the same algorithm. First the methylation in beta values were z-score standardized. The resulting matrix (X) of methylation z-scores (columns: CpG sites, rows: participants) was then multiplied with the transpose resulting in XX’. XX’ was then standardized by dividing the matrix with the mean of the diagonal, resulting in an average value of 1 for the diagonal of M. We used the R package BGData 2.1.0 (28) to compute the similarity matrix.

The next step is to regress the outcomes on M and covariates using a mixed effects model fitted with REML. Fixed effects covariates included several variables known to be associated with DNAm levels: sex, maternal age, maternal smoking, maternal education, cell type proportions, gestational age, birth weight (unless a variable was the outcome). M and batch were defined as random effects.

The average of multiple imputations was used to avoid potential bias due to missing data and to make analyses more comparable between outcomes by including the same set of participants. We used the covariate and outcome variables to predict missing variables in 100 imputations with 30 iterations using MICE (29) in R. Further analyses were then performed using the average value across the imputations, or the most often occurring category.

According to power analyses with genetic data, to accurately estimate the variance explained using GREML methods, large sample sizes are necessary. Especially with less heritable traits sample sizes above 5,000 participants are recommended (30). Currently, studies that have measured DNAm and child outcomes in more than 1000 participants are rare. While the power requirements for DNAm data are unclear, there is nevertheless a high risk of sampling variance, with results randomly changing heavily depending on a particular sample composition. We attempted to reduce these risks by estimating R^2^_Methylation_ in two independent cohorts, as well as by performing cross-validation within cohorts.

Cross-validation (CV) was applied in the following way: 1. M was estimated across all participants. 2. Eighty percent of the sample was randomly chosen as training sample and the GREML model was fitted in this training sample. 3. Based on the results of the training sample the best linear unbiased predictions (BLUP) were extracted for the test sample. The BLUP estimates reflect the extent to which participants are predicted to have above or below average outcome values, based on their similarity in genome-wide methylation to other participants. 4. The outcome is predicted based on M and the covariates 5. The predictions are correlated with the actual observed outcome and squared to obtain the variance explained by the model. 6. The variance explained by a covariate only model is subtracted to obtain the variance explained by DNAm beyond the other tested variables (ΔR^2^_Methylation_) 7. Step 1-6 are repeated to have results for 1000 random training-testing splits (Monte-Carlo cross-validation) 8. The mean estimate of ΔR^2^_Methylation_, with standard deviation across the cross-validation splits are extracted. 9. Results of both cohorts are averaged, weighted by the inverse of the cross-validation variance.

These analyses were run with the qgg package in R, which has implemented GREML models with cross-validation (31). We wrote additional functions, which can be found in the omicsR2 package: https://github.com/aneumann-science/omicsR2. The provided functions simplify the process of comparing the predictive performance of DNA methylation compared to a covariates-only baseline model.

## Results

DNAm explained 0 to 50% of the tested outcomes’ variances. See Table 2 for full results and Figure 1–4 for a graphical representation of the estimate distribution across cross-validations.

**Table 2:** Variance explained by genome-wide DNA methylation

| Outcome | Covariates | ALSPAC |  | Generation R |  | Pooled |  |
| --- | --- | --- | --- | --- | --- | --- | --- |
| | | $\Delta R^2_{\text{Methylation}}$ | $SD_{CV}$ | $\Delta R^2_{\text{Methylation}}$ | $SD_{CV}$ | $\Delta R^2_{\text{Methylation}}$ | $SD_{CV}$ |
| Gestational Age | Basic | 0.369<br>[0.169;0.556] | 0.103 | 0.649<br>[0.367;0.795] | 0.109 | 0.502<br>[0.184;0.783] | 0.075 |
|  | Full | 0.309<br>[0.137;0.475] | 0.089 | 0.592<br>[0.352;0.719] | 0.094 | 0.443<br>[0.163;0.704] | 0.065 |
| Birth Weight | Basic | 0.109<br>[-0.016;0.241] | 0.065 | 0.241<br>[0.073;0.398] | 0.082 | 0.159<br>[0.004;0.375] | 0.051 |
|  | Full | 0.079<br>[0.000;0.179] | 0.047 | 0.207<br>[0.077;0.334] | 0.065 | 0.122<br>[0.008;0.314] | 0.038 |
| BMI | Basic | -0.005<br>[-0.052;0.022] | 0.019 | 0.002<br>[-0.026;0.022] | 0.011 | 0.000<br>[-0.040;0.022] | 0.009 |
|  | Full | -0.005<br>[-0.037;0.008] | 0.012 | -0.001<br>[-0.010;0.002] | 0.004 | -0.001<br>[-0.026;0.004] | 0.004 |
| ADHD | Basic | 0.000<br>[-0.025;0.026] | 0.012 | 0.001<br>[-0.033;0.030] | 0.014 | 0.000<br>[-0.030;0.029] | 0.009 |
|  | Full | -0.001<br>[-0.024;0.012] | 0.008 | -0.001<br>[-0.020;0.010] | 0.008 | -0.001<br>[-0.022;0.010] | 0.006 |
| IQ | Basic | -0.001<br>[-0.037;0.020] | 0.013 | 0.000<br>[-0.004;0.008] | 0.005 | 0.000<br>[-0.024;0.018] | 0.004 |
|  | Full | -0.001<br>[-0.014;0.002] | 0.004 | -0.001<br>[-0.006;0.002] | 0.002 | -0.001<br>[-0.010;0.002] | 0.002 |

**Basic** sex and batch
$\Delta R^2_{\text{Methylation}}$ Variance explained by genome-wide DNA methylation minus variance explained by covariates [95% of values between lower;upper bound]
$SD_{CV}$ Standard-deviation of cross-validation estimates

**Figure 1:**
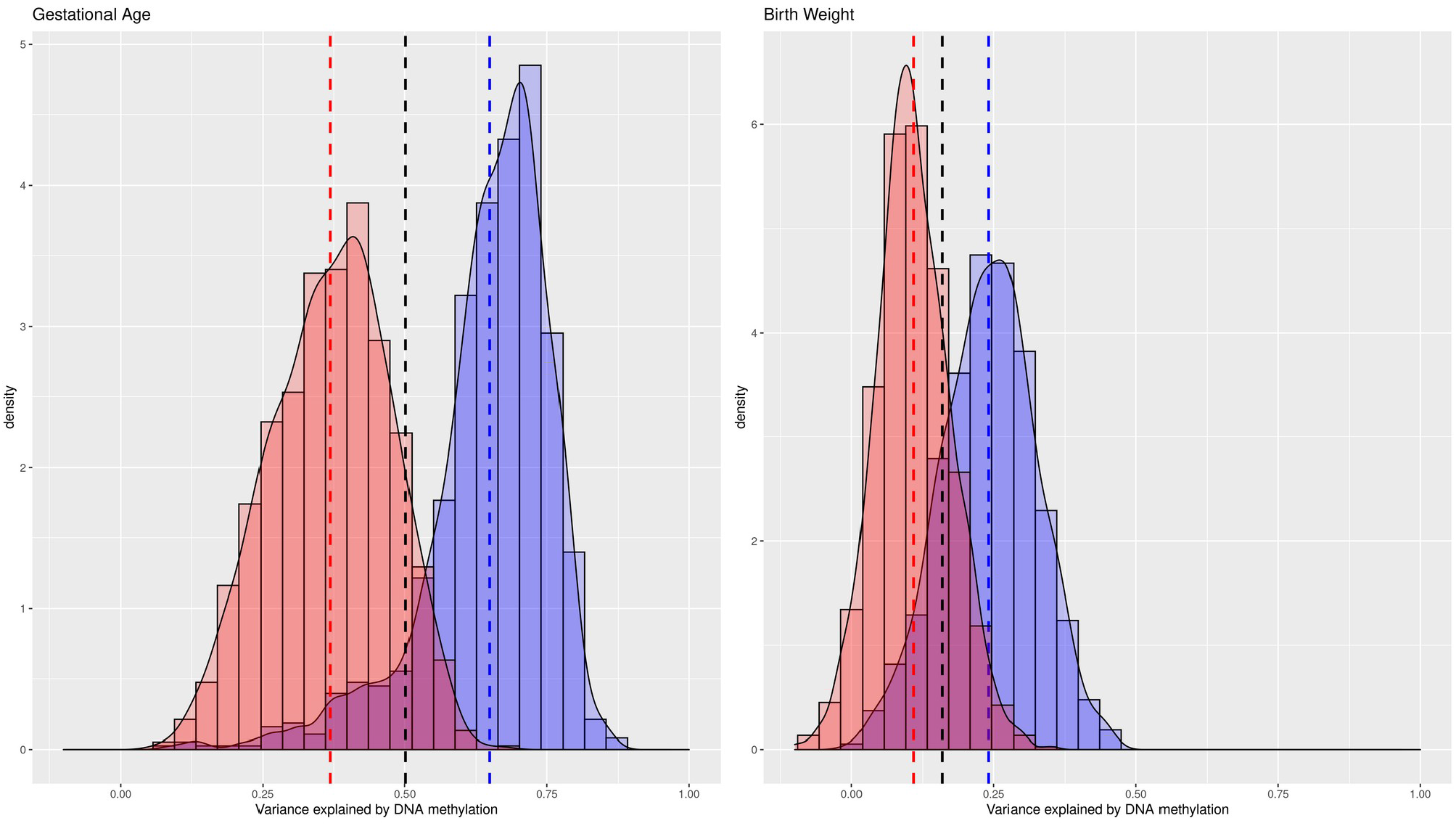
Variance explained in birth outcomes by cord blood DNA methylation (basic adjustment). Cross-validation distribution of ΔR^2^_Methylation,_ the variance explained by genome-wide DNA methylation minus variance explained by covariates (sex and batch) in ALSPAC (red) and Generation R (blue). Vertical lines indicate mean ΔR^2^_Methylation_ in ALSPAC (red), Generation R (blue) and a pooled estimate (black).

**Figure 2:**
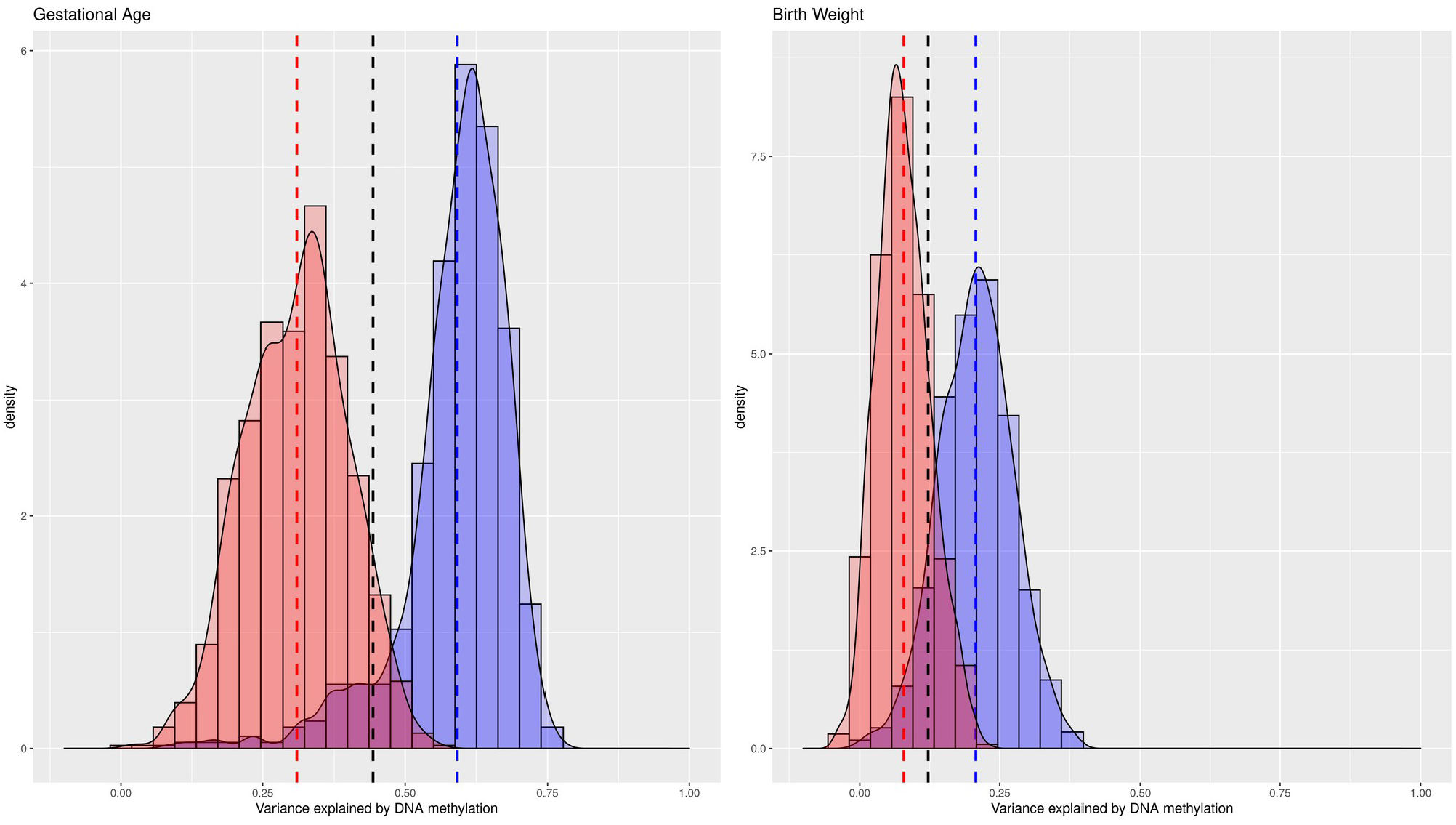
Variance explained in birth outcomes by cord blood DNA methylation (full adjustment). Cross-validation distribution of ΔR^2^_Methylation,_ the variance explained by genome-wide DNA methylation minus variance explained by covariates (sex, maternal age, maternal smoking, maternal education, cell type proportions, batch, gestational age*, birth weight* (* not when outcome is gestational age or birth weight)) in ALSPAC (red) and Generation R (blue). Vertical lines indicate mean ΔR^2^_Methylation_ in ALSPAC (red), Generation R (blue) and a pooled estimate (black).

**Figure 3:**
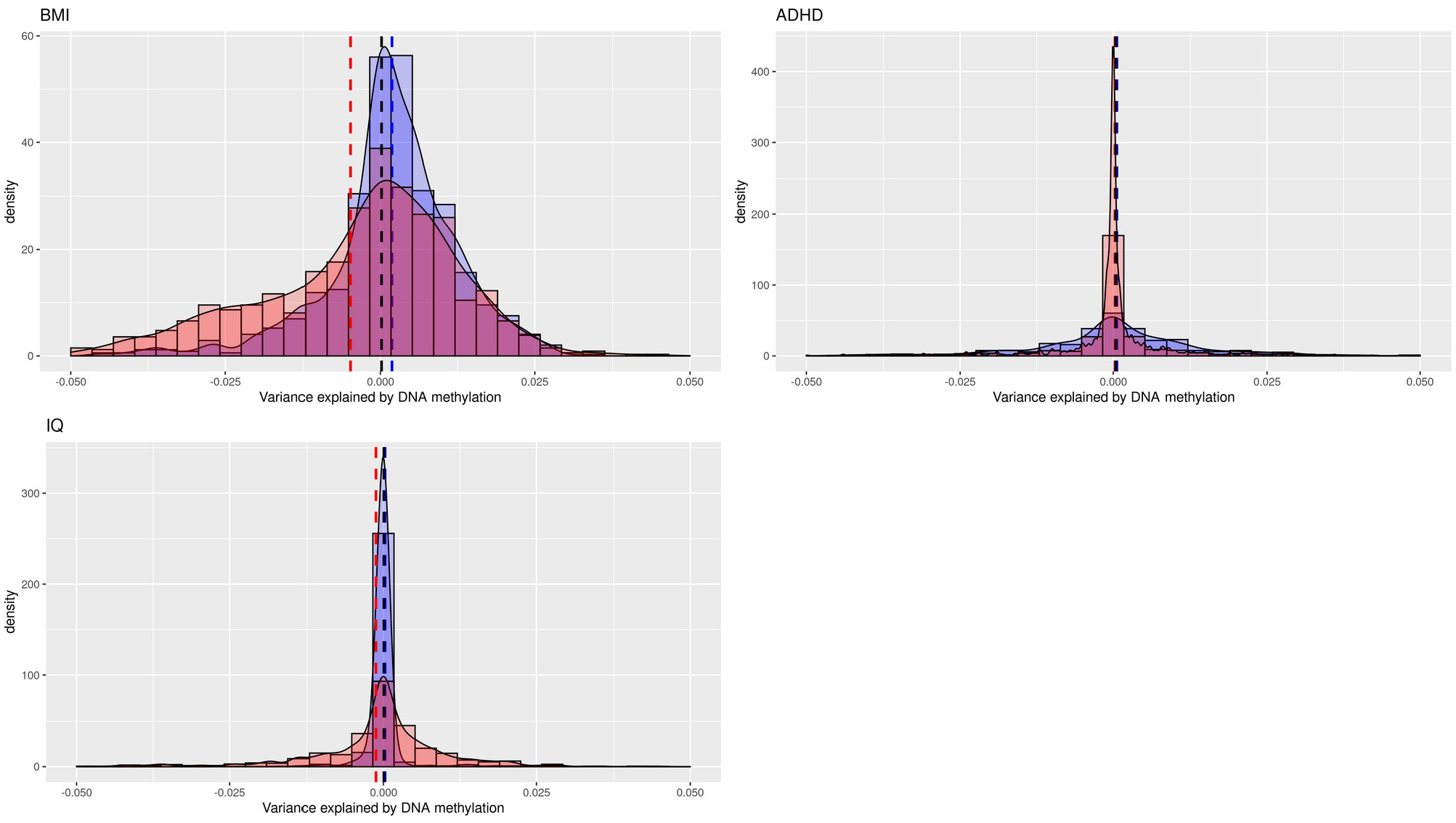
Variance explained in childhood outcomes by cord blood DNA methylation (basic adjustment). Cross-validation distribution of ΔR^2^_Methylation,_ the variance explained by genome-wide DNA methylation minus variance explained by covariates (sex and batch) in ALSPAC (red) and Generation R (blue). Vertical lines indicate mean ΔR^2^_Methylation_ in ALSPAC (red), Generation R (blue) and a pooled estimate (black).

**Figure 4:**
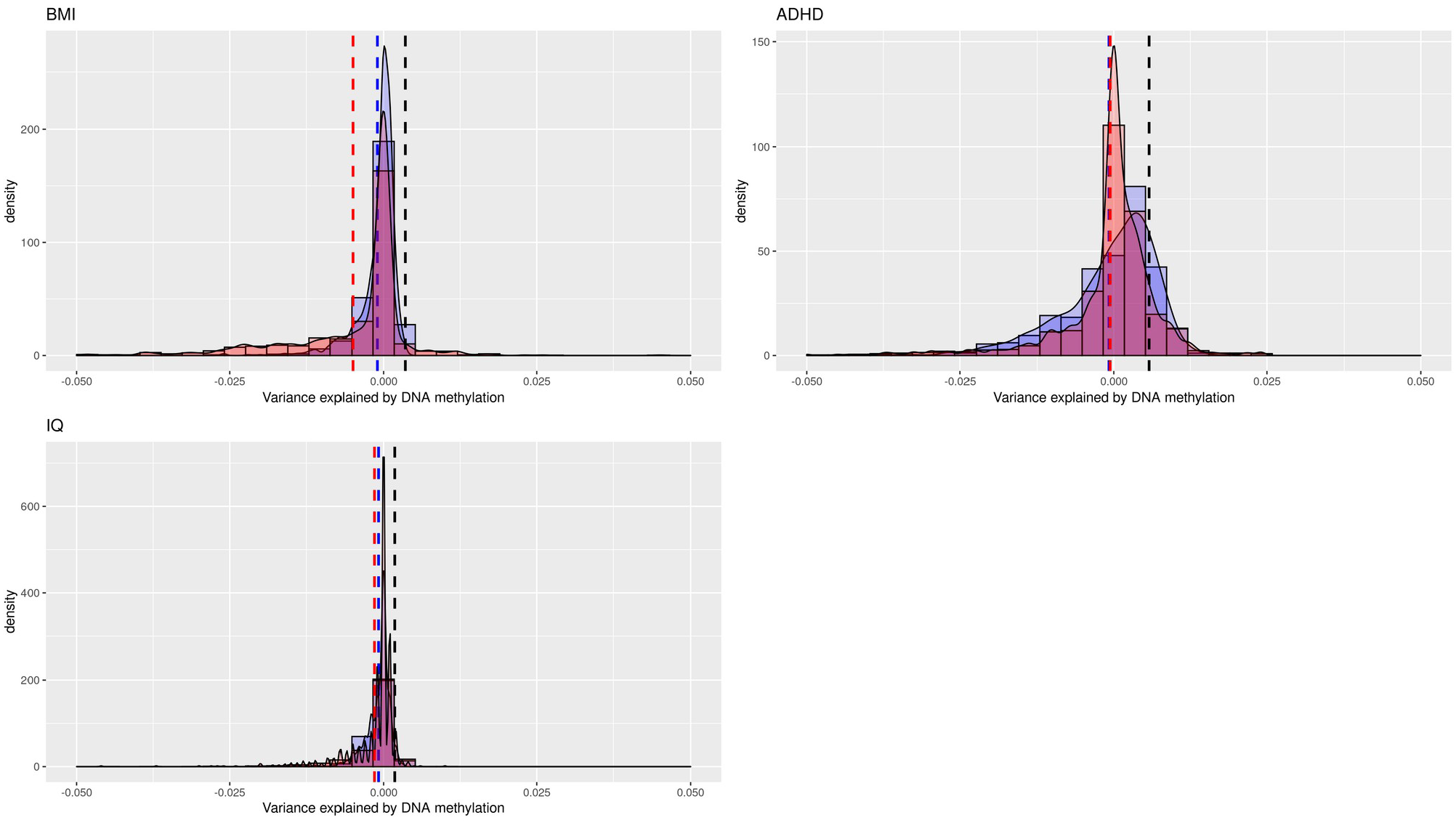
Variance explained in childhood outcomes by cord blood DNA methylation (full adjustment). Cross-validation distribution of ΔR^2^_Methylation,_ the variance explained by genome-wide DNA methylation minus variance explained by covariates (sex, maternal age, maternal smoking, maternal education, cell type proportions, batch, gestational age, birth weight) in ALSPAC (red) and Generation R (blue). Vertical lines indicate mean ΔR^2^_Methylation_ in ALSPAC (red), Generation R (blue) and a pooled estimate (black).

Gestational age had the highest R^2^ with 50.2% of the variance in gestational variance explained by DNAm in cord blood independent of sex and batch. In a fully adjusted model, 44.3% (SD_CV_ = 0.065) of variance was explained by DNAm. Notably, the ΔR^2^_Methylation_ was twice as large in GenR (ΔR^2^_Methylation_ = 59.2%, SD_CV_ = 0.094) compared to ALSPAC (ΔR^2^_Methylation =_ 30.9%, SD_CV_ = 0.089). Across both cohorts 95% of cross-validation results ranged from 16.3% to 70.4%, with 62.4% of values overlapping between the Generation R minimum and ALSPAC maximum.

For birth weight, the variance explained was estimated at 15.9% (SD_CV_ = 0.051) with basic adjustment and 12.2% (SD_CV_ = 0.038) with full covariate adjustment. Again, the estimate was much larger in Generation R (ΔR^2^_Methylation_ = 20.7%, SD_CV_ = 0.065) compared to ALSPAC (ΔR^2^_Methylation_ = 7.9%, SD_CV_ = 0.047). In the fully adjusted model, 95% of estimates were between 0.8% and 31.4% and most cross-validation estimates overlapped between these two cohorts (80.5%).

DNAm in cord blood did not explain variance in any of the childhood outcomes at school age (BMI, ADHD and IQ). This result was consistent in both cohorts, in which all cross-validation estimates were very close to 0, with the vast majority (97.5%) of estimates being below 2% in both basic and fully adjusted model. Correspondingly, the cross-validation standard deviations were below 0.1%, suggesting that no matter which participants were randomly assigned to training or validation, the estimated effect was always near 0.

## Discussion

This study is the first to report the extent to which childhood outcomes are explained by cord blood genome-wide DNAm. We observed that methylation patterns explained substantial variance for gestational age, moderate variance for birth weight and no variance explained for prospective associations with BMI, IQ or ADHD symptoms at school-age.

A strength of the study was the use of two cohorts, which are among the largest samples of cord blood methylation currently available. Both cohorts are comparable in many ways, for instance they represent populations of European ancestries living in western European countries and similar outcome assessment ages. In addition, cord blood DNAm assessment was very similar, as both cohorts used the same methylation array and were normalized jointly.

The general trend of results regarding ranking from highest to lowest explained outcomes agreed between the cohorts. The highest estimates across both cohorts were found for gestational age, which is consistent with previous studies. Bohlin et al. tested a prediction model based on 58-132 CpG sites in cord blood using similar covariates (sex, maternal age, maternal smoking, cell composition) as in our study (10). The authors were able to explain 50-65% of variance in a test sample of 685 participants from the MoBa cohort. Since we modeled a much higher number of probes, we would expect at least equal prediction performance in our study. The previous findings are consistent with the Generation R estimate of 59% variance explained and suggests that adding more probes from the Illumina 450k array would not increase performance of the prediction model.

However, the previous results are less consistent with the 31% estimate in ALSPAC, indicating either a higher variability in lower powered samples or a potential bias towards null effects in lower sample sizes, as we will discuss later. Another contributor to study heterogeneity may be the different methods used to estimate gestational age. Most gestational age estimates in ALSPAC were based on the last reported menstrual period, whereas in Generation R most estimates were based on ultrasound scans. The latter method is expected to have less measurement error and thus higher variance explained assuming constant methylation effects.

Genome-wide DNAm explained also explained variance in birth weight, albeit less so than for gestational age. Interestingly, the estimate was again higher in GenR than ALSPAC. In contrast to gestational age, there was no apparent noteworthy difference in birth weight assessment, yet the estimates differed even more between cohorts than for gestational age, so other potential causes for the observed study differences must be discussed. One cause could be higher sampling variance in lower sample sizes. The different estimates may hint that the ΔR^2^_Methylation_ values at sample sizes of around 1000 samples or lower may be highly variable, with lower sample sizes more likely to over or underestimate the true variance explained.

School-age outcomes showed a ΔR^2^_Methylation_ near zero for BMI, IQ and ADHD symptoms at age 6 in both cohorts. In contrast to gestational-age and birth weight, these analyses present prospective associations over at least 6 years and have resulted in fewer genome-wide significant findings in previous EWAS (13–15). This temporal component together with perhaps lower contribution of DNAm may weaken associations and result in lower variance explained estimates. While these factors lead to the expectation of a lower variance explained estimate in prospective estimates as opposed to cross-sectional analyses, estimates of 0% appear nevertheless unlikely. For instance, for ADHD, 9 CpG sites have been identified in a meta-analysis, in which most participants were drawn from ASLPAC and GenR (13). Both cohorts showed a high lambda in the EWAS, not accounted for by confounding, suggestive of a highly poly-epigenetic signal. Therefore, 0% variance explained estimates in a subset of the data is implausible. Besides a true lower variance explained for the school-age outcomes, a potential bias towards 0 values in underpowered samples may be at play as well.

Assuming a high uncertainty of ΔR^2^_Methylation_, we would expect a large standard deviation in the cross-validation distribution, as some iterations will randomly show a variance explained that is much too high or too low. However, in our study all analyses with outcomes showing a 0% ΔR^2^_Methylation_, had an estimate near 0% in almost all cross-validation iterations. This resulted in very small cross-validation standard deviations, much smaller compared to the gestational age or birth weight analysis. This is incompatible with a high estimate uncertainty due to low sample size. Hence, we suspect that a bias towards 0 estimates is at play if outcomes, which are not very strongly associated with DNAm, are analyzed in small samples. Such a behavior has been previously noted by GCTA author Jian Yang in the context of GREML when applied to genetic data (http://gcta.freeforums.net/thread/204/run-greml-analysis-small-sample). We therefore speculate that the true ΔR^2^_Methylation_ values for the school-age outcomes are likely to be higher than 0% and below estimates found for gestational age and birth weight, which themselves did not display a bias towards 0% estimates. Interestingly, early GCTA studies indicated no SNP heritability for child psychiatric phenotypes (32), but later larger multi-center GCTA (33), and LD-score regression studies (34) have since then repeatedly demonstrated a SNP heritable component. Contrary to genetic studies, an additional source of variability in DNA methylation is the assessment time point. Estimates for the school-age outcomes are likely different for concurrent DNA methylation measures than cord blood, but sample size was not sufficient for these analyses in the current study.

A limitation of the current analyses is the coverage of the 450k methylation array. The CpG sites measured by the array represent less than 2% of all CpG sites in the genome. While neighboring CpG sites tend to be correlated, CpG sites may also represent unmeasured CpG sites to a degree, but the correlations are not as stable or predictable as correlations between single nucleotide polymorphisms in linkage disequilibrium. Thus, the variance explained by array DNAm is unlikely the maximum which can be explained by all DNAm variation in humans. That said, the estimates do in theory represent the maximum that can be explained by the effects found in an EWAS using the same array, as it represents the joint effect of all measured CpG sites.

This study adjusted for a number of potential confounders, such as maternal smoking and education, as well as cell type proportions. Nevertheless, the observational nature of the study design makes it unclear whether the strong association between DNAm and gestational age represent direct effects of DNAm on gestational age, the effects of gestational age on DNAm, or the effect of unmeasured confounding. Furthermore, we only measured DNAm in a single tissue (cord blood). As DNAm can be tissue-specific, other tissue may show higher associations with studied outcomes, e.g. adipose tissue and body weight.

Despite the current limitations due to sample size, the results of the gestational age analysis demonstrate that GREML methods are applicable to studies of DNA methylation. We expect that increases in sample size will make this analytical approach more reliable for outcomes less strongly associated with DNAm. An increase in sample size would also allow for more complex questions to be answered. For example, as the method we utilized enables one to fit multiple similarity matrices, it is in principle possible to estimate ΔR^2^_Methylation_ adjusted for genetic effects or to estimate the genome-wide interaction between genetic and epigenetic effects. Answers to these questions would not only be helpful in further understanding of how DNAm relates to development and health, but would also inform the design of future EWAS. For instance, EWAS might need to model interactions between genetics and methylation levels, if interactions on a genome-wide level are substantial (35).

In summary, we showed that genome-wide DNAm in cord blood explains almost half of the variance in gestational age. DNAm was also associated to a lesser degree with birth weight. DNAm at birth, however, did not explain variance in child BMI, IQ and ADHD symptoms at school-age. The GREML approach holds promise for elucidating the relationship between genome-wide DNAm, child development and health outcomes, but increases in sample sizes are required to accurately estimate outcomes that are less strongly associated with DNAm and to explore more complex models, which can integrate different highly dimensional data.

## Data Availability

The datasets generated and analyzed during the current study are not publicly available to ensure participant privacy and compliance with Dutch and UK law, but are available on reasonable request. For Generation R data, please contact management and the corresponding author. For ALSPAC, please see http://www.bristol.ac.uk/alspac/researchers/ for instructions on data access.

https://generationr.nl/researchers/collaboration/

## List of abbreviations

(EWAS): Epigenome-wide association studies
(BMI): Body mass index
(CV): Cross-validation
(DNAm): DNA methylation
(M): Methylation similarity matrix
(G): Genetic relatedness matrix

## Declarations

### Ethics approval and consent to participate

Ethical approval for the study was obtained from the ALSPAC Ethics and Law Committee and the Local Research Ethics Committees. Consent for biological samples has been collected in accordance with the Human Tissue Act (2004). Informed consent for the use of data collected via questionnaires and clinics was obtained from participants following the recommendations of the ALSPAC Ethics and Law Committee at the time.

All parents gave informed consent for their children’s participation. The Generation R Study is conducted in accordance with the Declaration of Helsinki. Study protocols were approved by the Ethics Committee of Erasmus MC.

### Consent for publication

Not applicable

### Funding

#### ALSPAC

The UK Medical Research Council (MRC) and Wellcome (Grant ref: 102215/2/13/2) and the University of Bristol provide core support for ALSPAC. This publication is the work of the authors and EW will serve as guarantors for the contents of this paper. A comprehensive list of grants funding is available on the ALSPAC website (http://www.bristol.ac.uk/alspac/external/documents/grant-acknowledgements.pdf). Methylation data in the ALSPAC cohort were generated as part of the UK BBSRC funded (BB/I025751/1 and BB/I025263/1) Accessible Resource for Integrated Epigenomic Studies (ARIES, http://www.ariesepigenomics.org.uk). EW was partially funded by the Bath Institute for Mathematical Innovation. EW is also funded by the European Union’s Horizon 2020 research and innovation programme (grant nº 848158) and by CLOSER, whose mission is to maximise the use, value and impact of longitudinal studies. CLOSER was funded by the Economic and Social Research Council (ESRC) and the Medical Research Council (MRC) between 2012 and 2017. Its initial five year grant has since been extended to March 2021 by the ESRC (grant reference: ES/K000357/1). The funders took no role in the design, execution, analysis or interpretation of the data or in the writing up of the findings. www.closer.ac.uk.

#### GENR

The general design of the Generation R Study is made possible by financial support from Erasmus MC, University Medical Center Rotterdam, Erasmus University Rotterdam, the Netherlands Organization for Health Research and Development (ZonMw) and the Ministry of Health, Welfare and Sport. The EWAS data was funded by a grant from the Netherlands Genomics Initiative (NGI)/Netherlands Organisation for Scientific Research (NWO) Netherlands Consortium for Healthy Aging (NCHA; project nr. 050-060-810), by funds from the Genetic Laboratory of the Department of Internal Medicine, Erasmus MC, and by a grant from the National Institute of Child and Human Development (R01HD068437). A. Neumann and H. Tiemeier are supported by a grant of the Dutch Ministry of Education, Culture, and Science and the Netherlands Organization for Scientific Research (NWO grant No. 024.001.003, Consortium on Individual Development). A. Neumann is also supported by a Canadian Institutes of Health Research team grant. The work of H. Tiemeier is further supported by a NWO-VICI grant (NWO-ZonMW: 016.VICI.170.200). The work of CC has received funding from the European Union’s Horizon 2020 Research and Innovation Programme under the Marie Skłodowska-Curie grant agreement No 707404.This project received funding from the European Union’s Horizon 2020 research and innovation programme (733206, LifeCycle; 633595, DynaHEALTH; 848158, EarlyCause, 874739, LongITools)) and from the European Joint Programming Initiative “A Healthy Diet for a Healthy Life” (JPI HDHL, NutriPROGRAM project, ZonMw the Netherlands no.529051022 and PREcisE project ZonMw the Netherlands no.529051023).

### Contributions

AN, EW and CC developed the study design and drafted the manuscript. AN and EW performed statistical analysis on the GenR and ALSPAC data respectively and wrote the omicsR2 package. EW and CC supervised the study and share senior authorship. JFF,JBP and HT advised on research design and statistical analysis. JFF manages the DNA methylation data in GenR. VWJD is GenR director and oversaw data collection. All authors revised the manuscript critically.

## Acknowledgments

We thank all the children and families who took part in this study, as well as the support of general practitioners, hospitals, midwives and pharmacies.

## ALSPAC

We are extremely grateful to all the families who took part in this study, the midwives for their help in recruiting them, and the whole ALSPAC team, which includes interviewers, computer and laboratory technicians, clerical workers, research scientists, volunteers, managers, receptionists and nurses.

## GENR

The Generation R Study is conducted by Erasmus MC in close collaboration with the Erasmus University Rotterdam, Faculty of Social Sciences, the Municipal Health Service Rotterdam area, the Rotterdam Homecare Foundation, Rotterdam and the Stichting Trombosedienst & Artsenlaboratorium Rijnmond (STAR-MDC), Rotterdam. The generation and management of the Illumina 450K methylation array data (EWAS data) for the Generation R Study was executed by the Human Genotyping Facility of the Genetic Laboratory of the Department of Internal Medicine, Erasmus MC, the Netherlands. We thank Mr. Michael Verbiest, Ms. Mila Jhamai, Ms. Sarah Higgins, Mr. Marijn Verkerk and Dr. Lisette Stolk for their help in creating the EWAS database. We thank Dr. A.Teumer for his work on the quality control and normalization scripts.

